# Mapping the common barriers to optimal COPD care in high and middle-income countries: qualitative perspectives from clinicians

**DOI:** 10.1101/2023.11.13.23298474

**Authors:** Orjola Shahaj, Anne Meiwald, Krishnan Puri Sudhir, Rupert Gara-Adams, Peter Wark, Alexis Cazaux, Abelardo Elizondo Rios, Sergey Avdeev, Elisabeth J Adams

**Affiliations:** Aquarius Population Health, London, UK; John Hunter Hospital, Australia; Universidad Nacional de Cordoba, Argentina; Autonomous University of Nuevo Leon, Mexico; Sechenov First Moscow State Medical University, Russia

**Author notes:** Correspondence: Dr Elisabeth J Adams, Aquarius Population Health, Unit 29 Tileyard Studios, London N7 9AH, UK.

## Abstract

**Background:** Although predominantly preventable and treatable, chronic obstructive pulmonary disease (COPD) is a leading cause of death globally. Guidelines for managing the condition are widely available, yet COPD care remains suboptimal in many settings, including high and middle-income countries (HICs and MICs). Several approaches are used to diagnose and manage COPD, resulting in substantial variation in its care pathways. This study aimed to explore how barriers to optimal COPD care vary across HICs and MICs by identifying common and unique barriers to COPD care in six countries to inform global policy initiatives for better care while addressing specific challenges.

**Methods:** Based on international and national guidelines, we mapped COPD care pathways for Australia, Spain, Taiwan, Argentina, Mexico, and Russia. Country-specific pathways were populated with published epidemiological, health economic, and clinical data identified through a pragmatic literature review. Semi-structured interviews with 17 respiratory care clinicians further informed and validated the pathways, data inputs, and key issues arising in each country. Thematic content analysis was used to analyse common and unique barriers across countries.

**Results:** Six themes were common in most HICs and MICs: *“Challenges in COPD diagnosis”, “Strengthening the role of primary care”, “Fragmented healthcare systems and coordination challenges”, “Inadequate management of COPD exacerbations”; “Limited access to specialised care” and, “Impact of underfinanced and overloaded healthcare systems”*. One theme, *“Insurance coverage and reimbursement challenges”*, was more relevant for MICs.

HICs and MICs differ in patient and healthcare provider awareness, primary care involvement, spirometry access, and specialised care availability. Both face issues with healthcare fragmentation, guideline adherence, and COPD exacerbation management. MICs also grapple with resource limitations and healthcare infrastructure challenges.

**Conclusion:** Many challenges to COPD care are the same in both HICs and MICs, underscoring the pervasive nature of these issues. While country-specific issues require customised solutions, there are untapped possibilities for implementing global respiratory strategies that motivate countries to manage COPD effectively. In addition to healthcare system-level initiatives, there is a crucial need for political prioritisation of COPD to secure the essential resources it requires.

## Introduction

Chronic obstructive pulmonary disease (COPD) is a respiratory condition characterised by irreversible airflow limitation and symptoms such as dyspnoea, cough, and sputum production. Despite being preventable and treatable (1), according to the Global Burden of Disease Study, in 2019, COPD accounted for 3.3 million deaths, globally (2). COPD is already the third leading cause of death globally, which is seven years ahead of the World Health Organisation’s (WHO) prediction of 2030 (3). By 2060, deaths from COPD are expected to rise to 5.4 million (4).

The highest age-standardised COPD prevalence is in high-income countries (HICs), where it persists as a significant public health issue due to the considerable healthcare, economic and societal burden (2). It is also an important issue in upper middle-income countries (UMICs) which are expected to experience an increasing burden of this disease as their economies grow and populations age (5). Moreover, compared to HICs, UMICs may have more limited resources to respond to increased demands on their health systems, including fewer healthcare facilities and limited availability of essential medications and technologies (6).

The Global Initiative for Obstructive Lung Disease (GOLD) provides guidance on a universal and standardised approach to COPD care (4). Despite this guidance being widely available, COPD is often suboptimally managed in practice, and guidelines are not followed (7–9). This is linked to variation in how COPD care is delivered between settings (10–12) and leads to heterogeneity in patient outcomes and quality of life (QoL) (13).

Currently, no well-established and widely recognised method exists for mapping practical aspects of care delivery to identify deviations from the guideline provisions. One approach is reported by Meiwald et al. for COPD Evidenced Care Pathways, which collated and illustrated the steps in delivering COPD care across four healthcare settings (14). The Evidenced Care Pathways were developed based on clinical guidelines, peer-reviewed articles, and clinician perspectives, providing a qualitative assessment of differences between clinical practice and guidelines.

A granular understanding of barriers to providing guideline-adherent care in COPD and how they differ across countries with diverse healthcare systems could provide useful insights for policymakers and healthcare providers. The insight could improve policy planning, effective resource allocation and the successful implementation of initiatives to improve care (15).

This study seeks to identify common and unique barriers preventing optimal COPD care in three HICs (Australia, Spain and Taiwan) and three UMICs (Argentina, Mexico and Russia) to better inform global policy initiatives that tackle shared barriers (16) while ensuring that unique issues are addressed through more tailored approaches.

## Material and methods

This study used a previously reported mixed-method approach (14) to generate Evidenced Care Pathways. A qualitative analysis of the clinician interviews was conducted to evaluate each country’s themes. A narrative synthesis was used to compare themes in HICs and UMICs. The definition of HICs and UMICs was based on the World Bank classification (17).

### Development of Evidenced Care Pathways

Pragmatic searches were conducted on PubMed and Google Scholar to identify: 1) international and national guidelines for the diagnosis and management of COPD and 2) economic, epidemiological and clinical data about COPD in the study settings. See Supplement 1 for the search strategy. The literature review findings informed the pathway mapping and the discussion guides for the clinician interviews.

Initially, a generic pathway outlining the steps to COPD care was drafted for each country based on the GOLD recommendations. It was further refined using country-specific guidelines (18–23). The pathways were then used as a visual aid in the clinician interviews to assess how current care delivery deviates from guideline provisions. Further changes to the pathways to reflect current care delivery were made following clinicians’ input, using an iterative approach until the final structure of the Evidenced Care Pathway was achieved.

### Qualitative analysis of barriers to optimal COPD care

Semi-structured interviews with 17 clinicians were used to develop and validate the structure of the COPD Evidenced Care Pathways and understand clinician-perceived barriers to optimal COPD care. The qualitative analysis followed the consolidated criteria for reporting qualitative research (COREQ) checklist (24).

#### a) The research team and reflexivity

A summary of the characteristics of the research team and their relationship with the participants is provided in Table 1.

**Table 1.**
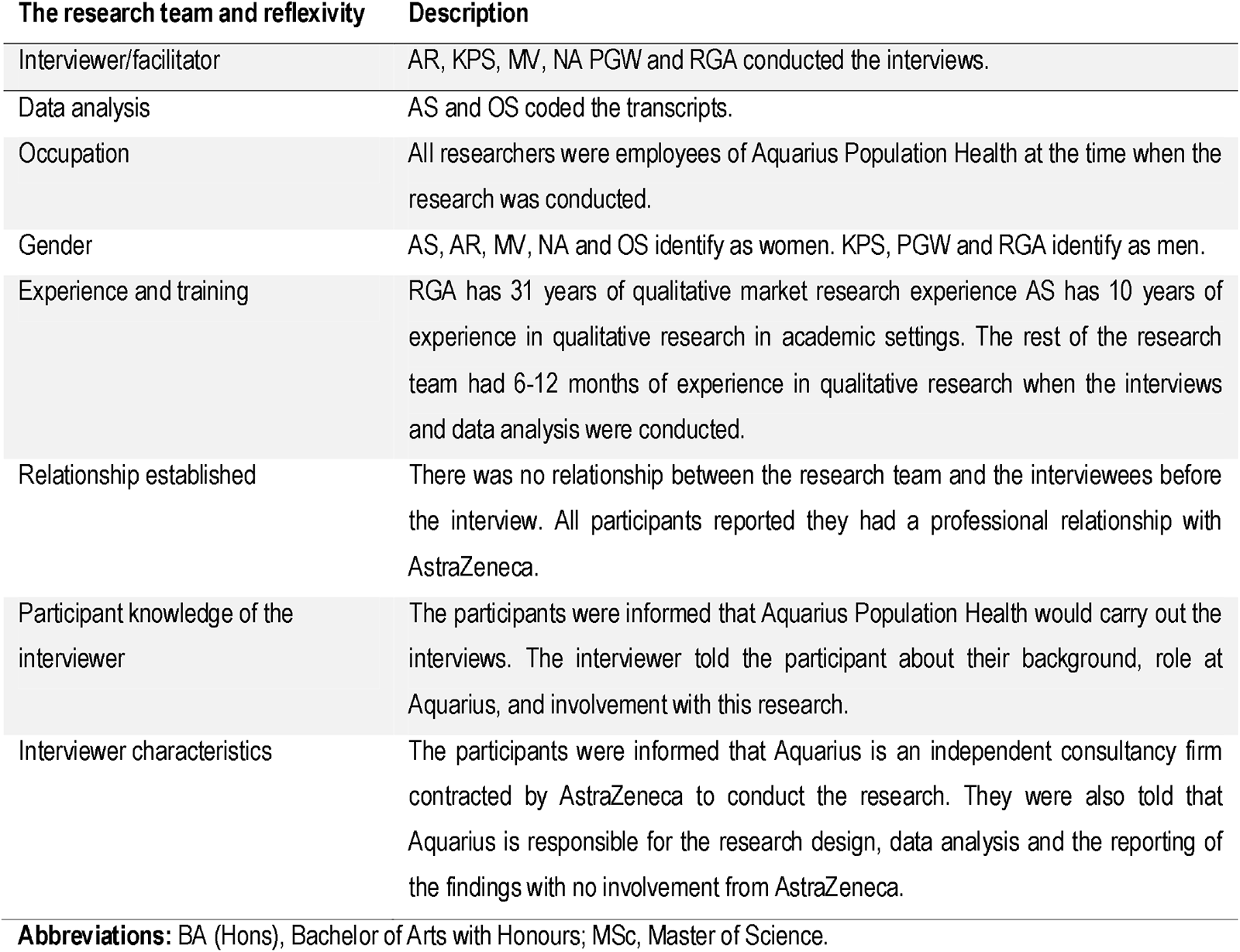
Characteristics of the research team and their connection with the relationship with the participants.

| The research team and reflexivity | Description |
| --- | --- |
| Interviewer/facilitator | AR, KPS, MV, NA PGW and RGA conducted the interviews. |
| Data analysis | AS and OS coded the transcripts. |
| Occupation | All researchers were employees of Aquarius Population Health at the time when the research was conducted. |
| Gender | AS, AR, MV, NA and OS identify as women. KPS, PGW and RGA identify as men. |
| Experience and training | RGA has 31 years of qualitative market research experience AS has 10 years of experience in qualitative research in academic settings. The rest of the research team had 6-12 months of experience in qualitative research when the interviews and data analysis were conducted. |
| Relationship established | There was no relationship between the research team and the interviewees before the interview. All participants reported they had a professional relationship with AstraZeneca. |
| Participant knowledge of the interviewer | The participants were informed that Aquarius Population Health would carry out the interviews. The interviewer told the participant about their background, role at Aquarius, and involvement with this research. |
| Interviewer characteristics | The participants were informed that Aquarius is an independent consultancy firm contracted by AstraZeneca to conduct the research. They were also told that Aquarius is responsible for the research design, data analysis and the reporting of the findings with no involvement from AstraZeneca. |

**Abbreviations:** BA (Hons), Bachelor of Arts with Honours; MSc, Master of Science.

#### b) Participant selection

A total of 39 potential participants were approached through an email invitation, and 17 agreed to participate in the study. The participants met the following criteria: 1) healthcare professionals (HCPs) working in Argentina, Australia, Mexico, Spain, Russia, or Taiwan at the time of the study; and 2) who played an active role in diagnosing or managing COPD in any healthcare setting (primary, secondary or tertiary). Purposive sampling was used in all the countries. The choice of the countries was guided by the need to ensure diversity in geography and healthcare systems.

#### c) Data collection

Interviews took place between April – September 2021 online via Microsoft Teams . Consent to participate, record and transcribe the interview was obtained in advance from all participants via email. Participants were informed of the estimated length of the discussion. An example of an interview discussion guide is provided in Supplement 2. Two interviewers were present in each call; a lead and a notetaker. An interpreter was present in three interviews. In these cases, only the English translation was transcribed. Seven interviews were conducted in Spanish (both interviewer and interviewee were native speakers), and certified translators later translated the transcripts into English for the thematic analysis. Transcripts were not returned to participants for correction. All participants consented to receive follow-up emails with clarifying questions regarding the Evidenced Care Pathways and were asked to validate the final structure. They were offered the option to receive an honorarium at the end of the process.

#### d) Data analysis

The interview data were analysed using deductive and inductive thematic analysis (25). Transcripts were coded using an existing framework (14). Data within each theme was then coded line by line to create sub-categories (inductive component). Particular attention has been paid to identifying both common and unique barriers. A note was made if the subtheme implied some form of improvement in COPD care. AS coded the transcripts. OS double-coded 20% of the data (four transcripts) (Table 1). Discrepancies were resolved through discussion. Results were anonymised before reporting.

A narrative synthesis was deployed to compare the themes and subthemes across the countries. A note was made where themes or subthemes were not applicable to a country and details of the country-specific nuances were captured separately.

## Ethics

No personal or patient-specific information was collected during the interviews. Participants were asked questions regarding the healthcare services for COPD in their country. Therefore, the project was deemed a service evaluation, and no ethics approval or review was necessary. Any data collected before, during and after the interview was handled following the European Union’s General Data Protection Regulation (GDPR) legislation.

## Results

### Evidenced Care Pathways

The detailed Evidence Care Pathways for Argentina, Australia, Mexico, Spain, Russia and Taiwan are presented in Supplement 2. Figure 1 shows an example of the overview page for an Evidenced Care Pathway. The pathways for all countries showed consistency in the organisation and care delivery for COPD. They split across patient touchpoints with the healthcare system over three key components: pre-diagnosis, diagnosis, and management.

**Figure 1.**
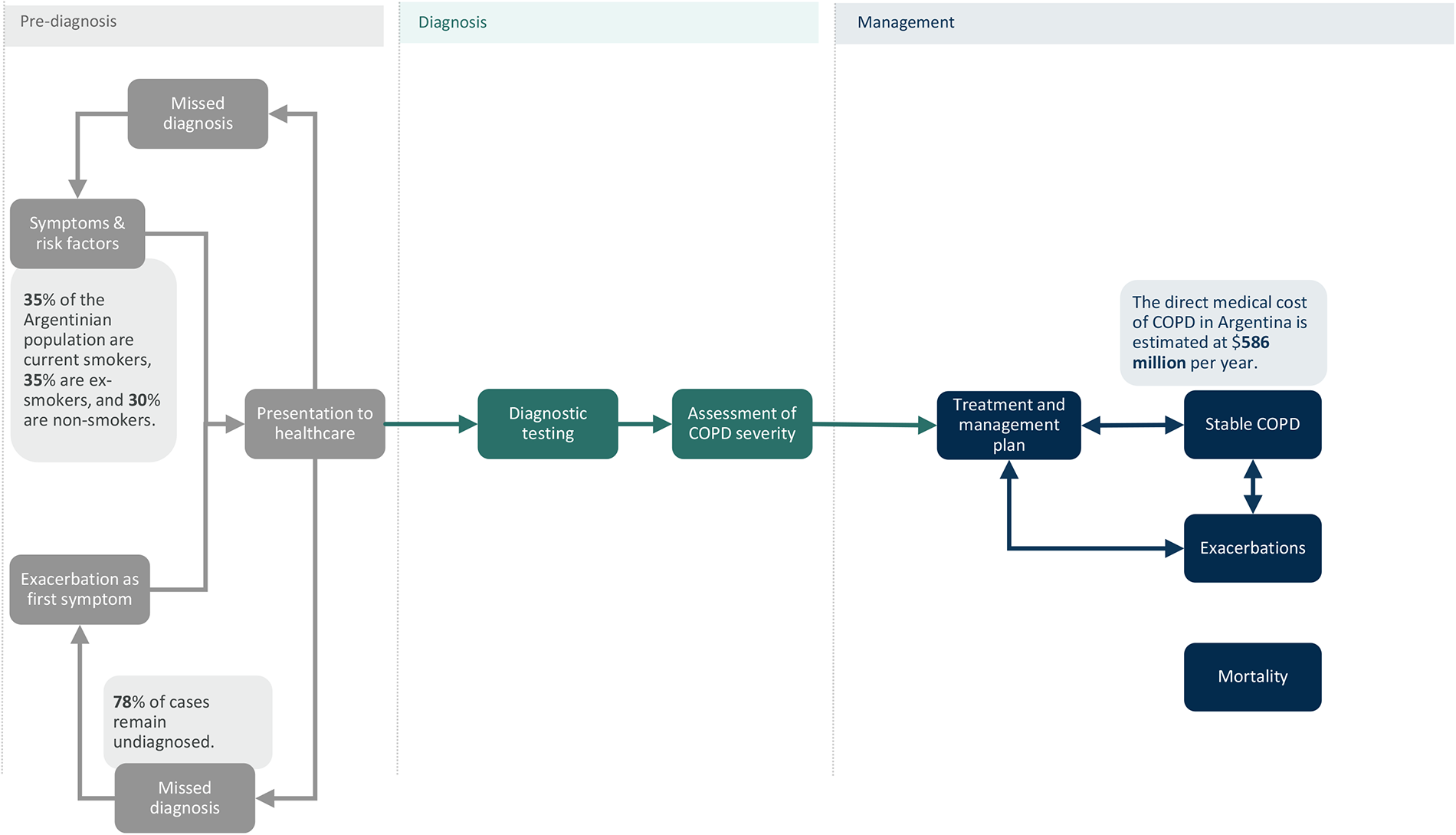
Top-level presentation of the Evidenced Care Pathway. Example from the Argentinian pathway.

However, differences between the Evidenced Care Pathways were observed. These are mainly related to how patients access specialist care, the tests used for COPD diagnosis, differences in the treatment algorithms, guidelines for managing COPD, and treatment reimbursement.

### Themes from qualitative analysis of interviews

Seven themes were identified, covering barriers to COPD care across the six countries. Table 2 lists the universal primary themes, level two sub-themes, and quotes supporting them. Table 3 shows where there was variation between countries for the primary themes and pinpoints if there were country-specific nuances within the subthemes.

**Table 2.**
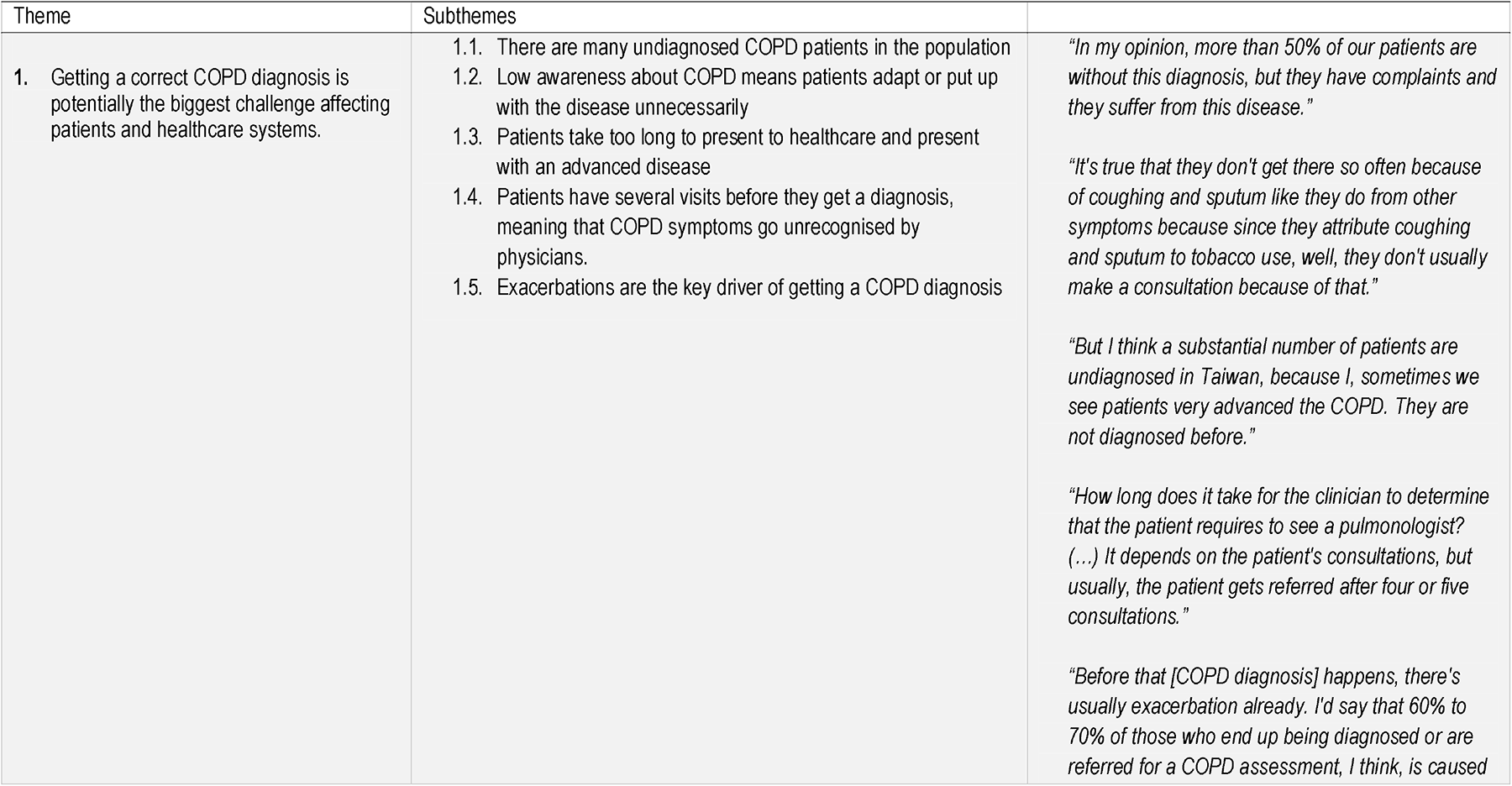
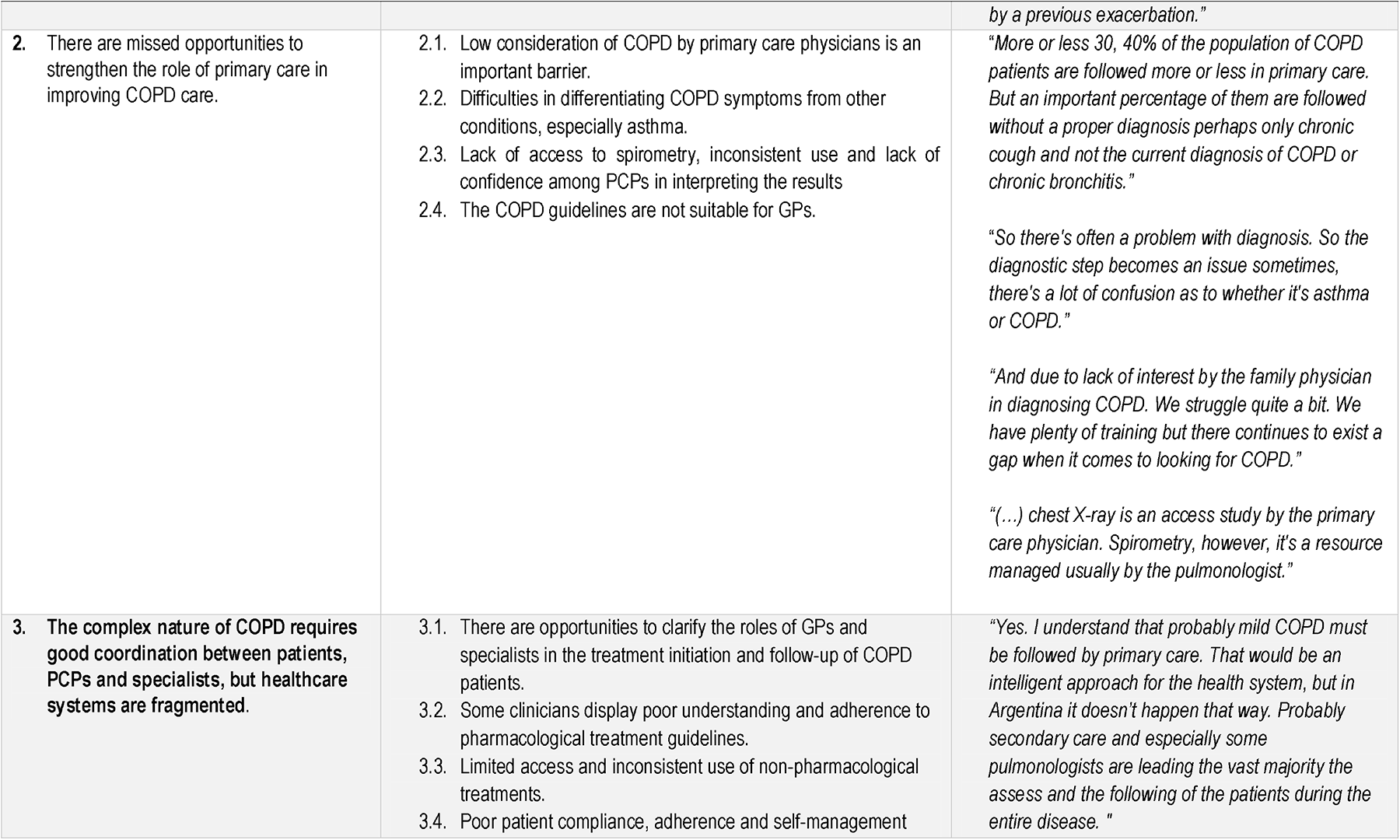
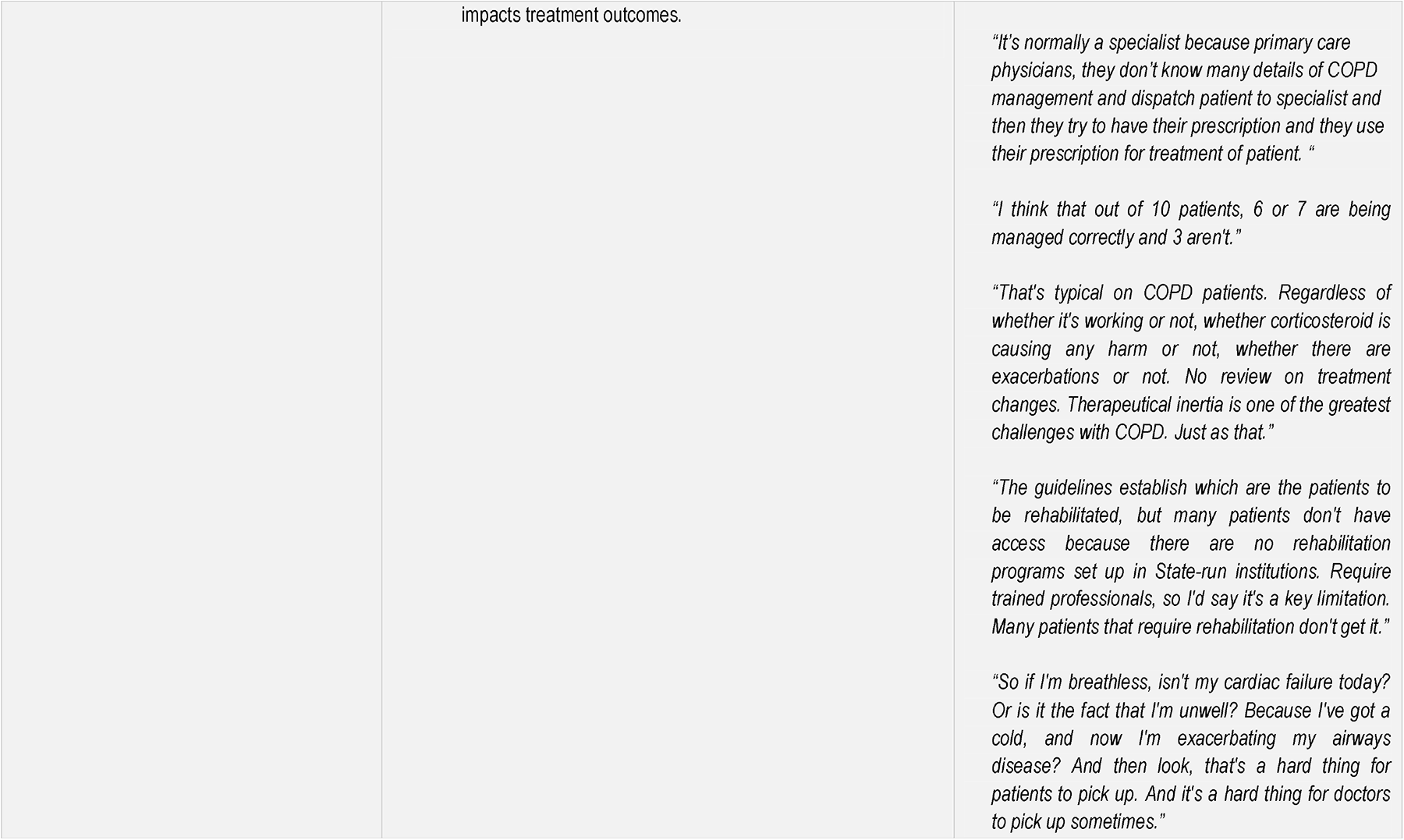
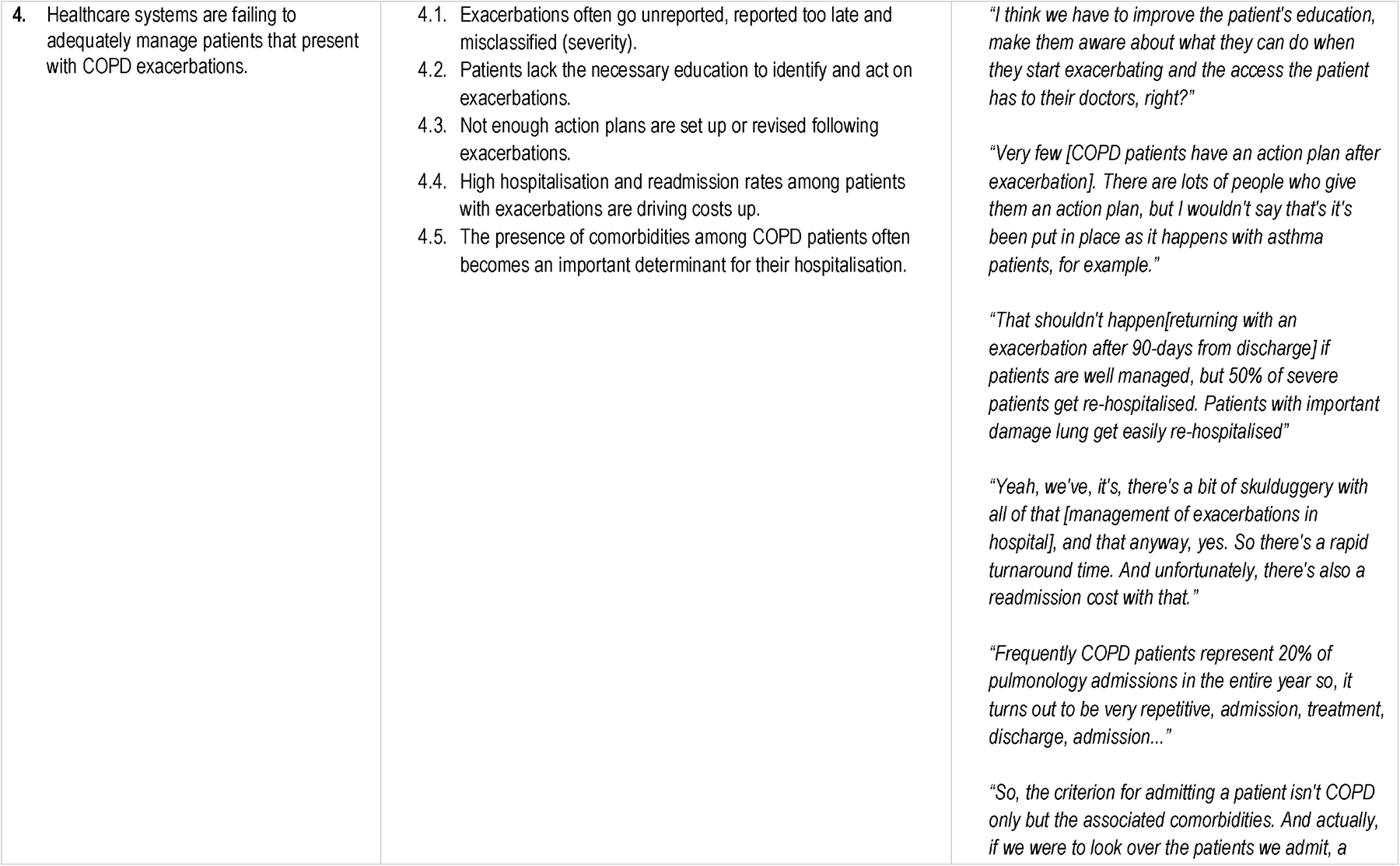
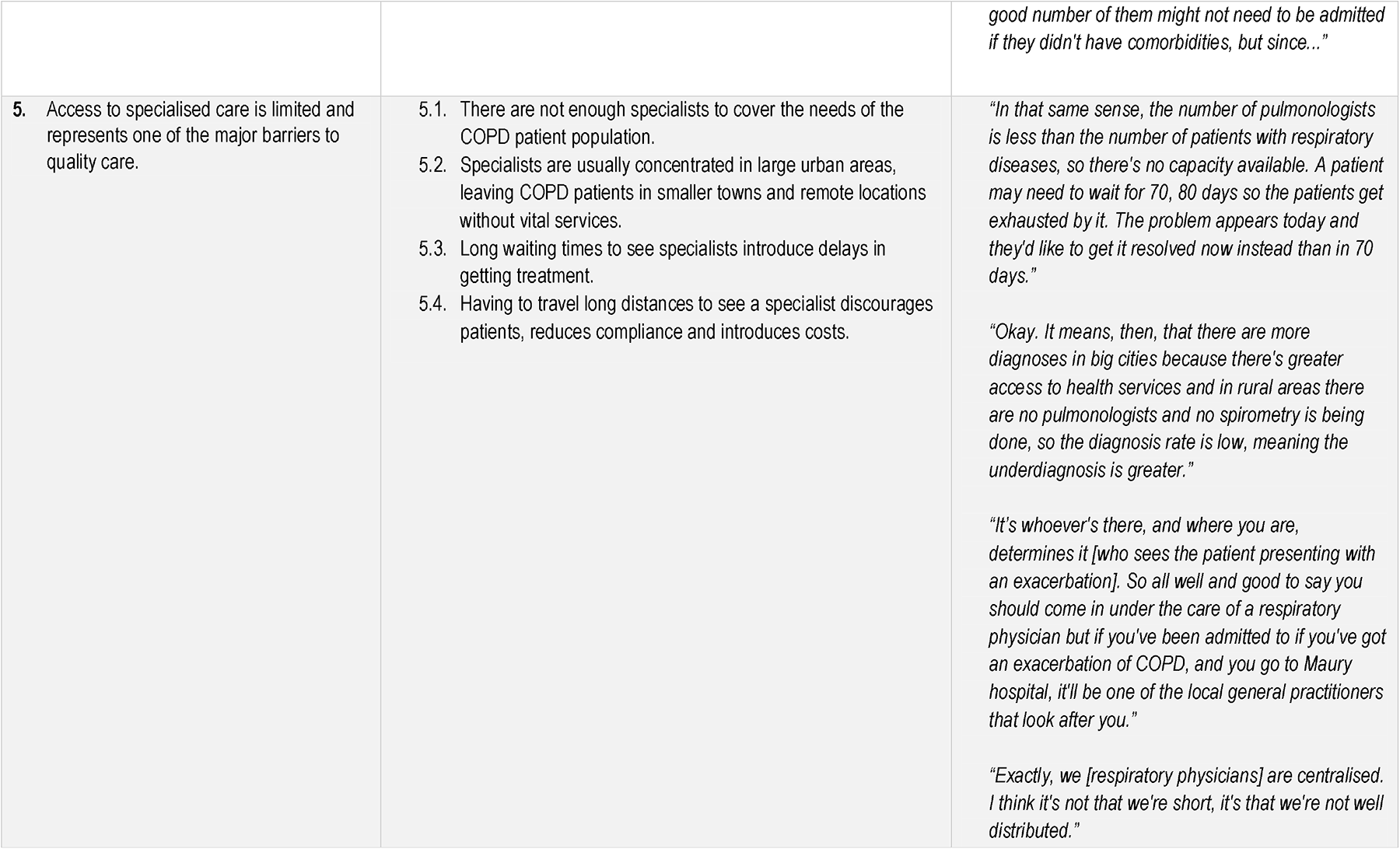
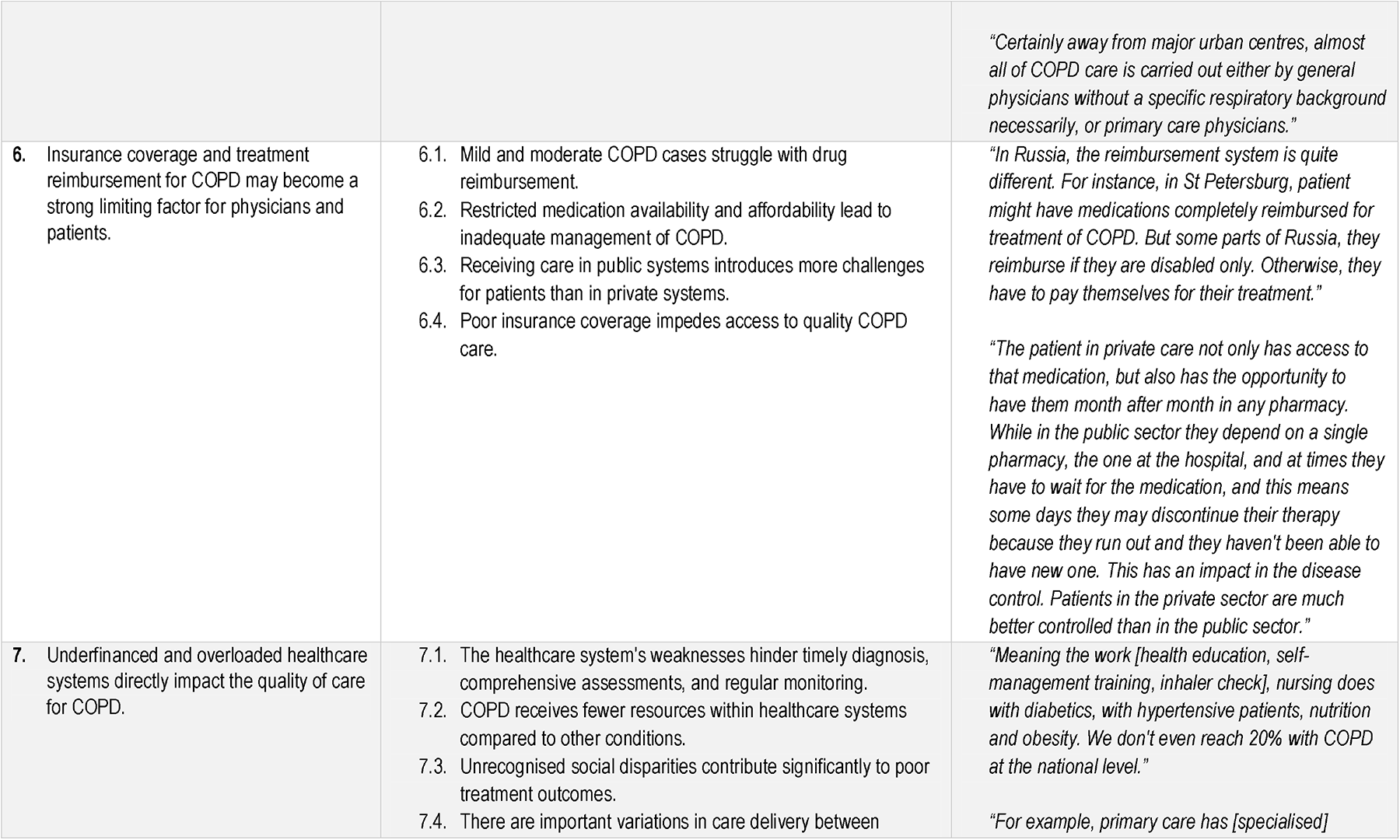
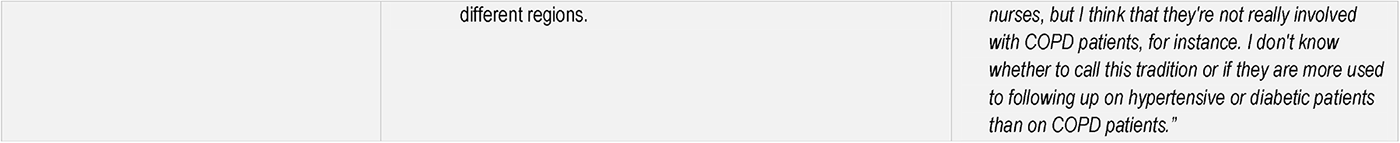
Themes, subthemes and quotes identified from analysing 17 interview transcripts in six countries: Argentina, Australia, Mexico, Russia, Spain, and Taiwan.

**Table 3.** The variation in primary themes between countries and indications where country-specific nuances were identified at the subtheme level.

| Themes and subthemes |
| --- |
| **Theme 1: Getting a correct COPD diagnosis is potentially the biggest challenge affecting patients and healthcare systems.** |
| 1.1 There are many undiagnosed COPD patients in the population. |
| 1.2 Low awareness about COPD means patients adapt or put up with the disease unnecessarily. |
| 1.3 Patients take too long to present to healthcare and present with advanced disease. |
| 1.4 Patients have several visits before they get a diagnosis, meaning that COPD symptoms go unrecognised by physicians. |
| 1.5 Exacerbations are the key driver of getting a COPD diagnosis. |
| **Theme 2: There are missed opportunities to strengthen the role of primary care in improving COPD care.** |
| 2.1 Low consideration of COPD by primary care physicians is an important barrier. |
| 2.2 Difficulties in differentiating COPD symptoms from other conditions, especially asthma. |
| 2.3 Lack of access to spirometry, inconsistent use and lack of confidence among PCPs in interpreting the results. |
| 2.4 The COPD guidelines are not suitable for GPs. |
| **Theme 3: The complex nature of COPD requires good coordination between patients, PCPs and specialists, but healthcare systems are fragmented.** |
| 3.1 There are opportunities to clarify the roles of GPs and specialists in the treatment initiation and follow-up of COPD patients. |
| 3.2 Poor understanding and adherence to pharmacological treatment guidelines |
| 3.3 Limited access and inconsistent use of non-pharmacological treatments. |
| 3.4 Poor patient compliance, adherence and self-management impacts treatment outcomes. |
| **Theme 4: The complex nature of COPD requires good coordination between patients, PCPs and specialists, but healthcare systems are fragmented.** |
| 4.1 Exacerbations often go unreported, reported too late and misclassified (severity). |
| 4.2 Patients lack the necessary education to identify and act on exacerbations. |
| 4.3 Not enough action plans are set up or revised following exacerbations. |
| 4.4 High hospitalisation and readmission rates among patients with exacerbations are driving costs up. |
| 4.5 The presence of comorbidities among COPD patients often becomes an important determinant for their hospitalisation. |
| **Theme 5: Access to specialised care is limited and represents one of the biggest barriers to quality care.** |
| 5.1 There are not enough specialists to cover the needs of the COPD patient population. |
| 5.2 Specialists are usually concentrated in large urban areas, leaving COPD patients in smaller towns and remote locations without vital services. |
| 5.3 Long waiting times to see specialists introduce delays in getting treatment. |
| 5.4 Having to travel long distances to see a specialist discourages patients, reduces compliance and introduces costs. |
| **Theme 6: Insurance coverage and treatment reimbursement for COPD may become a strong limiting factor for physicians and patients.** |
| 6.1 Mild and moderate COPD cases struggle with drug reimbursement. |
| 6.2 Restricted medication availability and affordability lead to inadequate management of COPD. |
| 6.3 Receiving care in public systems introduces more challenges for patients than in private systems. |
| 6.4 Poor insurance coverage impedes access to quality COPD care. |
| **Theme 7: Underfinanced and overloaded healthcare systems directly impact the quality of care for COPD.** |
| 7.1 The healthcare system's weaknesses hinder timely diagnosis, comprehensive assessments, and regular monitoring. |
| 7.2 COPD receives fewer resources within healthcare systems compared to other chronic conditions. |
| 7.3 Unrecognised social disparities contribute significantly to poor treatment outcomes. |
| 7.4 There are important variations in care delivery between different regions. |

Indicates there are some country-specific nuances for the corresponding subtheme where a particular challenge or barrier for that subtheme is brought up by the clinicians.
Indicates there are some country-specific nuances for the corresponding subtheme with a positive nature where clinicians have noticed an improvement for that specific issue.
Indicates the primary theme and subthemes that do not apply to the selected countries.

Tables 4–10 provide detail on the unique features observed in some settings within a given theme.

**Table 4.** Country-specific nuances within Theme 1, Challenges in COPD Diagnosis.

|  |
| --- |
| **Country-specific features:**<br>• In Russia, Argentina, and Mexico, clinicians estimated that over 50% of COPD diagnoses are made after an exacerbation.<br>• In Spain, Taiwan, and Australia, clinicians thought this problem existed on a smaller scale (10-20%).<br>• In Russia, there was a perception that COPD prevalence is increasing due to the high prevalence of smoking across the country.<br>• In Argentina and Mexico, biomass was mentioned as an important risk factor for rural communities in addition to smoking.<br>• In Australia and Taiwan, clinicians believe that COPD awareness among patients and healthcare providers has improved in the last decade. |

**Table 5.**
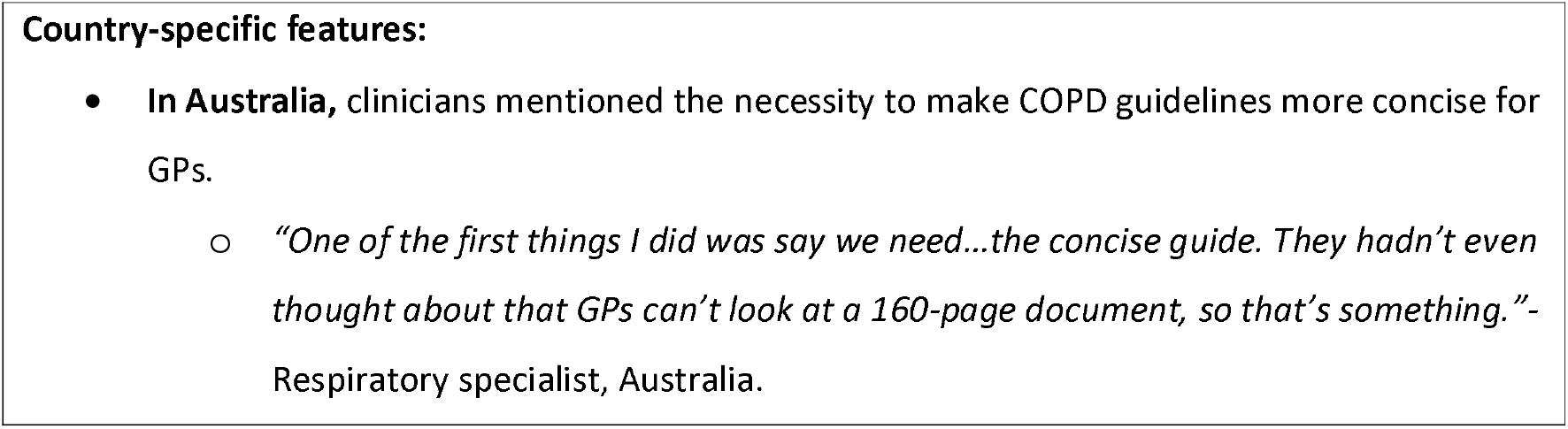
Country-specific features within Theme 2, Strengthening Primary Care in COPD Care.

|  |
| --- |
| **Country-specific features:**<br>• **In Australia**, clinicians mentioned the necessity to make COPD guidelines more concise for GPs.<br>○ *“One of the first things I did was say we need...the concise guide. They hadn’t even thought about that GPs can’t look at a 160-page document, so that’s something.”- Respiratory specialist, Australia.* |

**Table 6.**
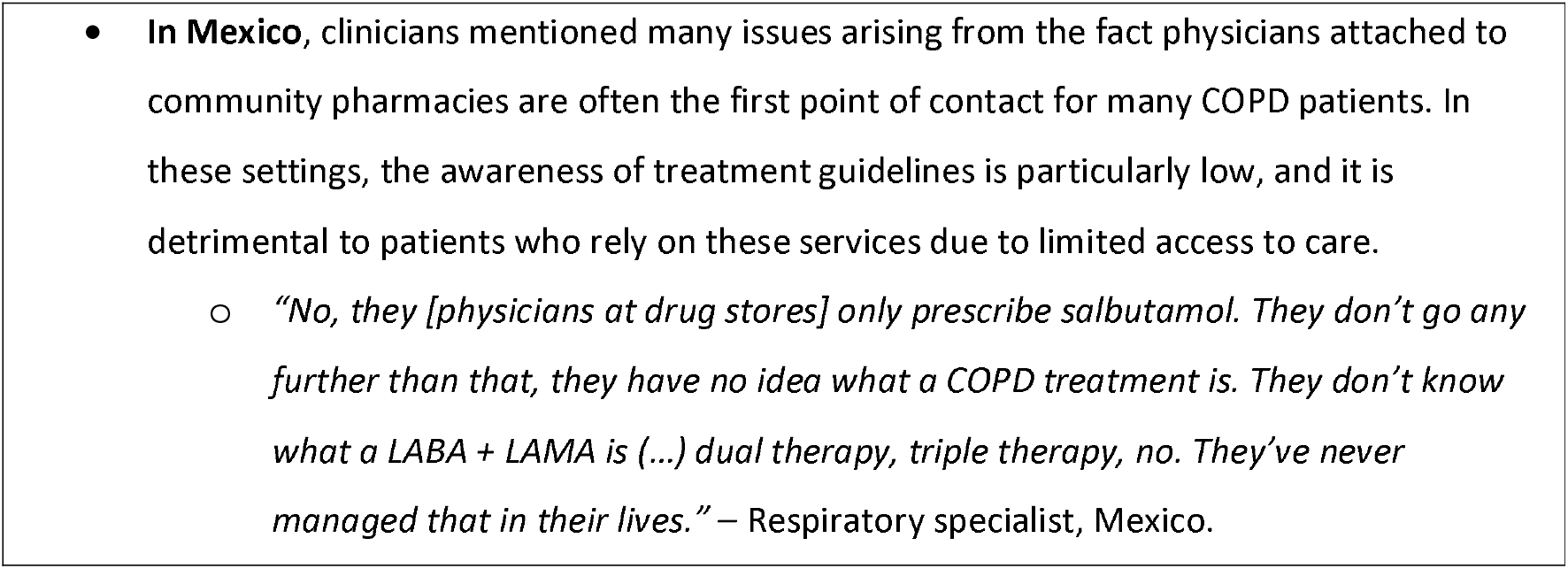
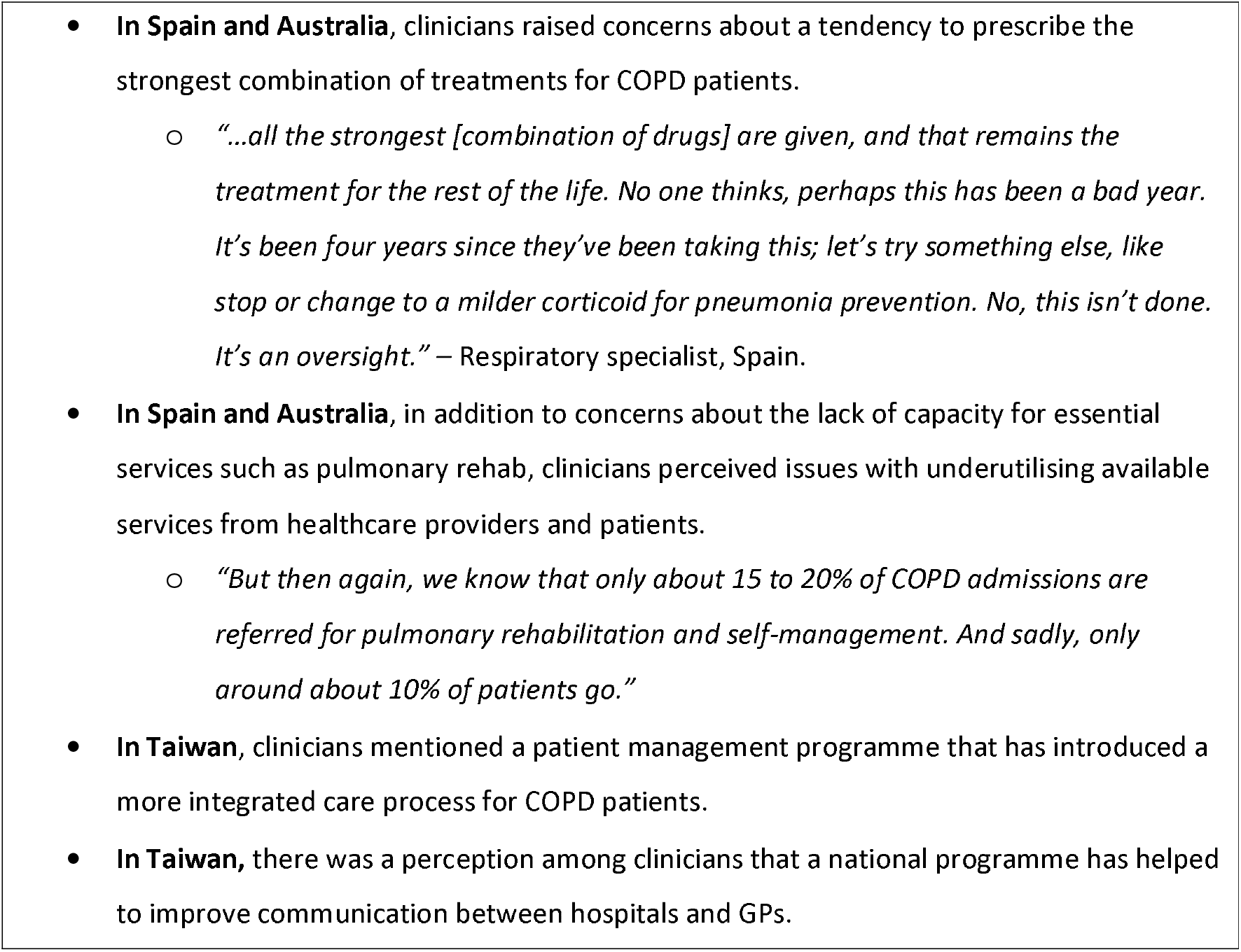
Country-specific features within Theme 3, Fragmented healthcare systems and coordination challenges.

**Table 7.**
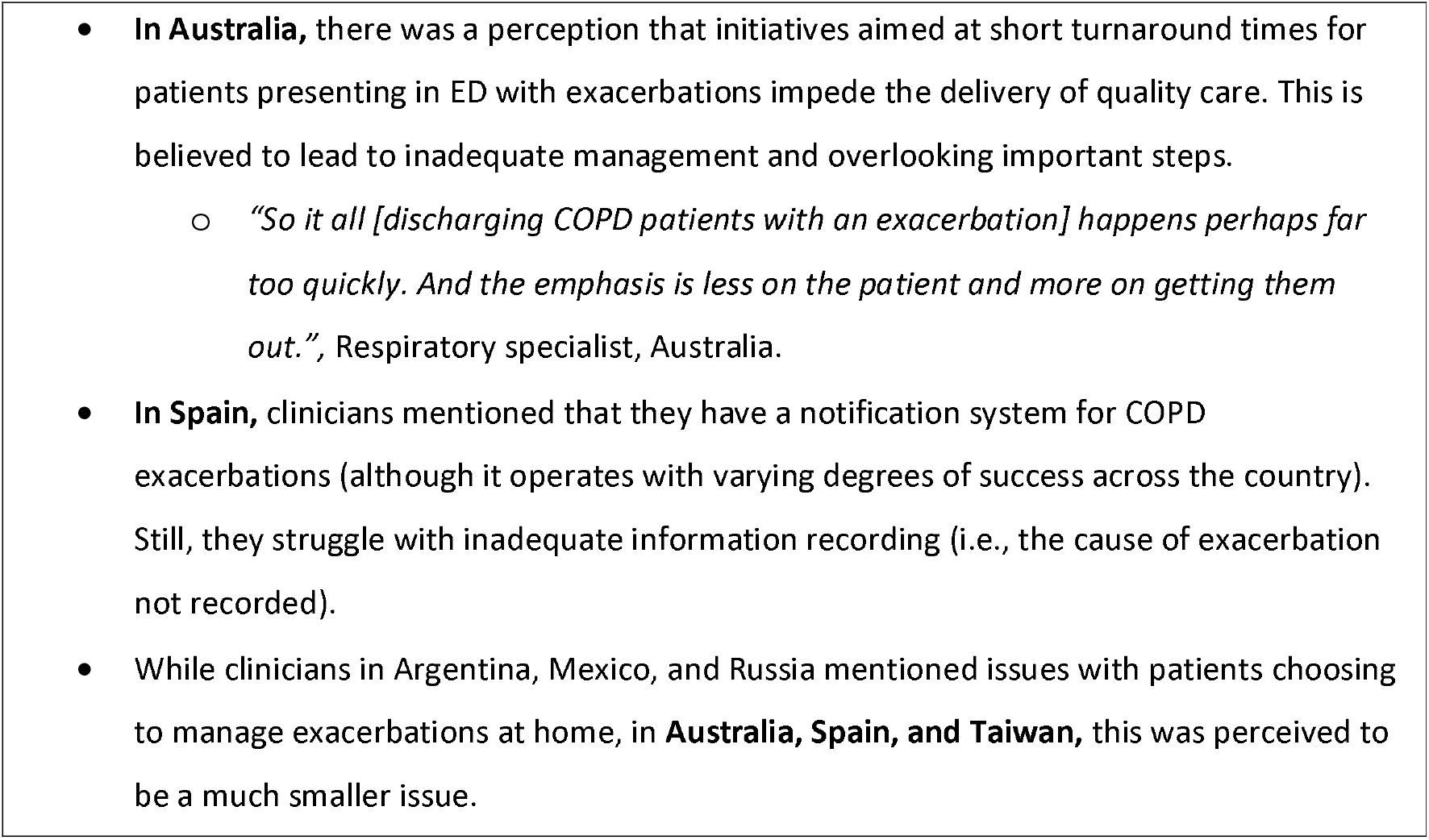
Country-specific features within Theme 4, Inadequate Management of COPD Exacerbations.

**Table 8.**
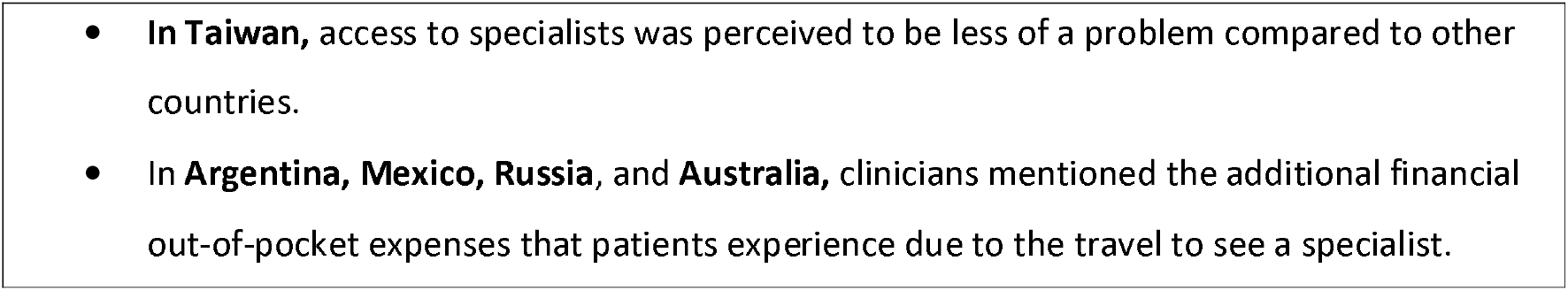
Country-specific features within Theme 5, Limited access to specialised care.

**Table 9.**
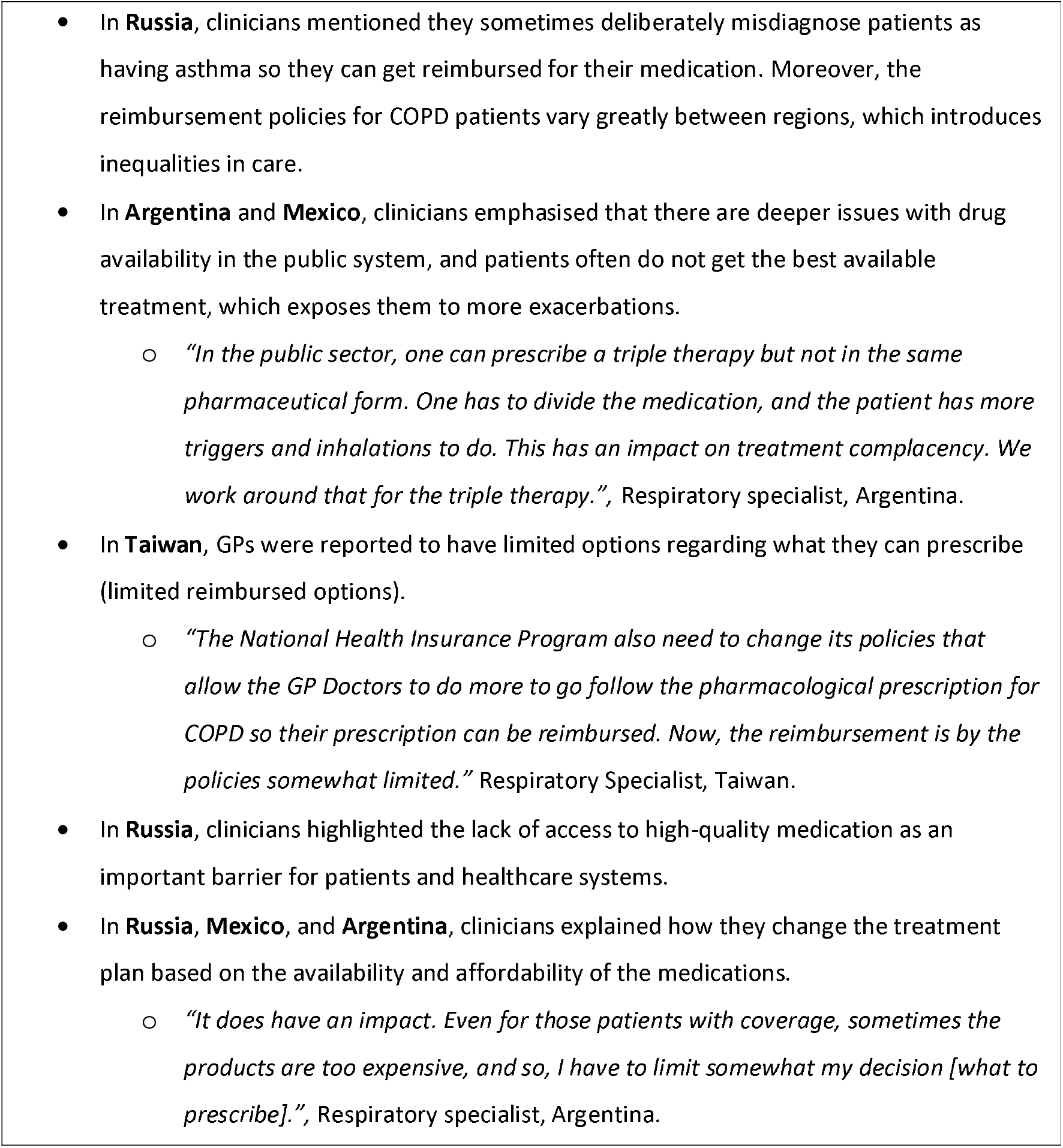
Country-specific features within Theme 6, Insurance coverage and reimbursement challenges.

**Table 10.**
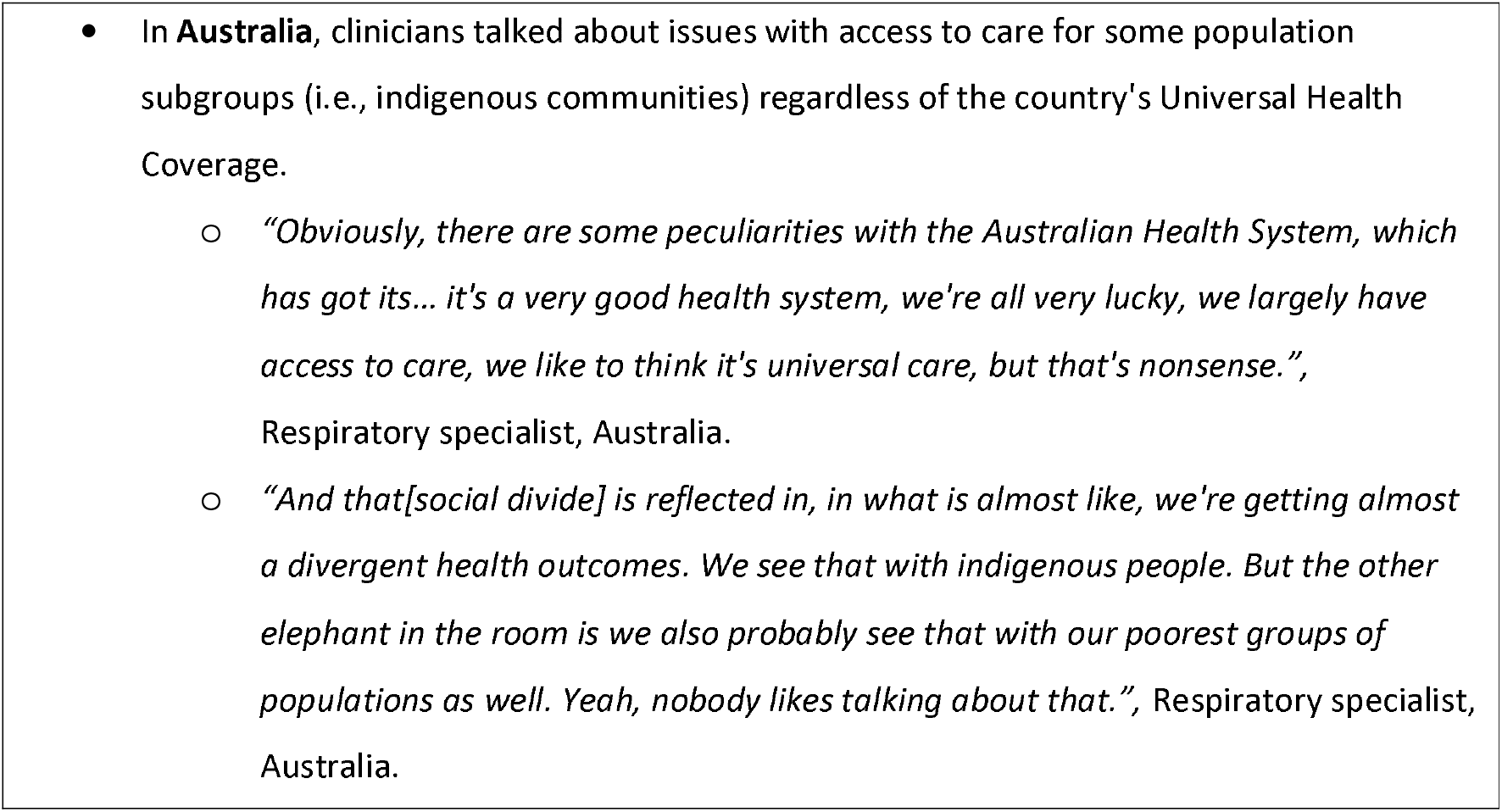
Country-specific features within Theme 7, Impact of underfinanced and overloaded healthcare systems.

#### Theme 1: Challenges in COPD diagnosis

Clinicians identified COPD diagnosis as a significant challenge affecting both patients and healthcare systems. Not having a COPD diagnosis hinders patients from entering the diagnostic pathway, leading to prolonged periods without treatment. Clinicians expressed concerns about the high prevalence of undiagnosed COPD cases, particularly in countries with a high prevalence of smoking or exposure to biomass fuels, such as Russia, Argentina, and Mexico.

Low awareness about COPD among patients was mentioned as a key factor leading them to adapt to or tolerate the disease without seeking proper medical attention. Clinicians observed that patients often presented with advanced stages of the disease, suggesting reasons such as delays in seeking healthcare, failure to consider COPD by clinicians, lack of spirometry to diagnose it and the lack of timely interventions.

> *“But I think a substantial number of patients are undiagnosed in Taiwan, because (…) sometimes we see patients very advanced the COPD. (sic) They are not diagnosed before.”* – Respiratory Specialist, Taiwan.

They also noted that COPD symptoms frequently went unrecognised during multiple visits to healthcare providers before a formal diagnosis was made. Most importantly, clinicians explained how exacerbations are often the driver for initiating the diagnosis process.

#### Theme 2: Strengthening the role of primary care in COPD

Clinicians emphasised the missed opportunities to enhance the role of primary care in improving COPD diagnosis. They expressed concerns about the limited consideration of COPD by primary care physicians (PCPs), who are typically patients’ first point of contact. Difficulties differentiating COPD symptoms from other conditions, particularly asthma, were highlighted as contributing to misdiagnosis and underdiagnosis.

> *“They even forget how [to interpret] lung function test[s] because they are more familiar with diabetes, more familiar with hypertension (…)”*, Respiratory Specialist, Taiwan.

Clinicians also identified the lack of access and inconsistent use to spirometry, and PCPs’ lack of confidence in interpreting results as significant barriers to timely COPD diagnosis.

Inconsistent use of spirometry in primary care settings adds to the diagnostic challenges, and subsequently, important bottlenecks are created in the system if patients rely only on respiratory specialists for diagnosis (Theme 3 and 4).

#### Theme 3: Fragmented healthcare systems and coordination challenges add to the complexity of managing COPD

COPD management depends on various factors, including interactions between healthcare providers and patients, healthcare infrastructure availability, and access to treatment options. The healthcare services that COPD patients need are divided between primary care, specialists (in secondary care) and emergency departments. Clinicians reported that there is some disconnect between these services, often leaving patients without adequate management.

Clinicians emphasised the need for effective coordination between patients, primary care physicians, and specialists in managing the complex nature of COPD. They recognised opportunities for clarifying the roles of general practitioners (GPs) and specialists in treatment initiation and follow-up. GPs’ lack of confidence and training in initiating treatment for mild cases was highlighted, reflecting missed opportunities for alleviating the excess burden on secondary care and reducing waiting times, which are important determinants in the patient journey (Theme 2, 5 and 7).

Poor understanding and adherence to pharmacological treatment guidelines were highlighted (mainly among PCPs but in some cases among specialists too). Clinicians expressed concerns about the inadequate understanding of treatment options among healthcare providers, leading to underusing or misusing pharmacological and non-pharmacological interventions.

> *“70% of [of COPD patients] in Mexico aren’t receiving adequate treatment. Because there’s steroid overuse. Things are the other way around here… A COPD patient is given a bunch of steroids, and an asthma patient is given a bunch of SABA. Things are upside down.”* Respiratory Specialist, Mexico.

They acknowledged the impact of poor patient compliance, adherence, and self-management on treatment outcomes which they believed is sometimes linked to insufficient resources dedicated to patient education. Substantial variation in guideline adherence was reported, particularly among GPs (Theme 1, subtheme 4).

#### Theme 4: Inadequate management of COPD exacerbations

Clinicians recognised the challenges in effectively managing COPD exacerbations. They observed frequent underreporting, delayed reporting, and misclassification of exacerbations.

> *“I think that up to 40% of the exacerbations don’t even get to be seen by a specialised physician or at the emergency care unit..”*, Respiratory specialist, Argentina.

Inadequate notification systems and lack of information sharing between healthcare providers reportedly contributed to the ineffective management of exacerbations.

> *“We are now doing some steps in this direction [setting up a notification system for exacerbations], but usually we know about the situation from our patient.”* – Respiratory Specialist, Russia.

Clinicians repeatedly emphasised the importance of patient education to enable early identification and appropriate response to exacerbations. They highlighted the need for action plans to be promptly set up or revised following exacerbations to optimise treatment outcomes. Discharged patients may not receive treatment reviews, further perpetuating the cycle of poor management.

Following discharge, patients reportedly need to wait a long time to see a specialist that can review their treatment plan. Clinicians mentioned all the above lead to high readmission rates, which drives costs up and increases pressure on healthcare systems. Comorbidities among COPD patients were also noted as an important determinant of hospitalisation, even in mild cases that otherwise would not have led to hospitalisation.

#### Theme 5: Limited access to specialised care

Clinicians highlighted limited access to specialised care as a significant barrier to quality COPD care. They expressed concerns about the shortage of specialists to meet the demands of the COPD patient population, resulting in long waiting times for consultations. This is linked to the over-reliance on specialists to diagnose and manage COPD patients (Theme 2). This theme did not apply to Taiwan, which has generally good access to specialist care.

The concentration of specialists in urban areas was identified as leaving COPD patients in smaller towns and remote locations without access to vital services. Clinicians noted that patients from rural or remote settings have to travel long distances to see a specialist. They felt this discouraged these patients from seeking care, reduced compliance, and introduced financial burdens.

> *“It’s because the number of specialists [in the big cities] is higher than in the rest parts of Russia (sic) historically. I believe that patients might have access to more types of treatment in these two cities [Moscow and St Peterburg] compared to other parts of Russia.”* Respiratory Specialist, Russia.

Clinicians mentioned that the increased waiting times to see a specialist can be particularly detrimental to COPD patients because they are either not receiving any treatment or they remain on the wrong treatment regimen, leaving patients at a higher likelihood of experiencing exacerbations.

> *“I would say if I refer the patient into our public outpatient, I would be warning the patient that it could be anywhere between three to six months before they’ll get seen.”* GP, Australia.

#### Theme 6: Insurance coverage and reimbursement challenges

This theme applies to Argentina, Mexico, and Russia.

Clinicians highlighted the impact of insurance coverage and reimbursement on COPD management. They observed challenges in drug reimbursement for mild and moderate COPD cases, as coverage often focuses on severe cases. In Russia, deliberate misdiagnosis (i.e. asthma) reportedly occurs to ensure reimbursement.

Restricted availability and affordability of medications were recognised as factors directly influencing clinical decision-making and leading to inadequate disease management. Moreover, in some countries clinicians emphasised the challenges patients’ face in public systems compared to private systems, and the negative impact of poor insurance coverage on access to quality COPD care.

#### Theme 7: Impact of underfinanced and overloaded healthcare systems

Clinicians identified underfinanced and overloaded healthcare systems as directly impacting the quality of COPD care. They highlighted system weaknesses that hindered timely diagnosis, comprehensive assessments, and regular monitoring of COPD patients. One example they stated is that short appointments do not allow for a thorough assessment, inhaler technique check, patient education, etc.

> *“Usually, a physician has to do that [inhaler technique check] during the patient visit. But as far as we know, sometimes the physician is very pressed for time during the visit and, unfortunately, this very important part of the consultation is absent.”*, Respiratory specialist, Russia.

Clinicians observed that COPD receives fewer resources within healthcare systems compared to other conditions, such as diabetes or cardiovascular diseases, indicating a need for increased attention and resource allocation. One frequently cited example was the need to have more COPD-dedicated specialist nurses.

> *““For example, primary care has [specialised] nurses, but I think that they’re not really involved with COPD patients, for instance. I don’t know whether to call this tradition or if they are more used to following up on hypertensive or diabetic patients than on COPD patients.”*, GP, Australia.

Even when the clinicians’ understanding of the available treatment options is sufficient, they reported often having restrictions regarding what they can prescribe or where to refer patients. They mentioned the lack of essential services such as pulmonary rehabilitation, smoking cessation, counselling, patient education, etc. Furthermore, affordability issues were seen to push clinicians to alter treatment plans based on reimbursement options rather than guideline-based care (Theme 6).

The contribution of unrecognised social disparities to poor treatment outcomes among certain COPD patients was emphasised. Addressing these disparities and promoting equity in access to care were identified as crucial steps to enhance COPD management. Variations between regions, particularly in countries with extensive geographical spreads, were noted.

## Discussion

### Main findings

This research indicated that barriers to optimal care for COPD patients are very similar across all countries, with five out of the seven themes identified in this research applying to all of them: *Challenges in COPD diagnosis; Strengthening the role of primary care; Fragmented healthcare systems and coordination challenges; Inadequate management of COPD exacerbations; and, Impact of underfinanced and overloaded healthcare systems*. This finding implies that the challenges faced in delivering effective COPD care are not limited to specific countries and are shared across different contexts. The presence of similar barriers suggests that these issues are pervasive and need to be addressed globally to improve COPD management. It also highlights that COPD continues to be under-prioritised, irrespective of a country’s economic development status and the resilience of its healthcare systems.

Only one theme (*Insurance coverage and reimbursement challenges*; Theme 6) was reportedly more relevant to the UMICs included in this study (Argentina, Mexico and Russia) and was not reported as a barrier by clinicians in HICs (Australia, Spain and Taiwan). Another difference was that Theme 5, *Limited access to specialised care*, did not apply to Taiwan, where patients have better access to specialists and have short waiting times to see them (26).

The development of the Evidenced Care Pathways offered a clear and concise overview of how COPD care is currently delivered across six diverse healthcare settings, depicting the complex interplay of various stakeholders, interventions, and transitions involved in the care process.

### Comparing the findings with previous similar work

Although broadly similar, the theme structure identified in this research differs from the one reported by Meiwald et al. (14), where only four HICs were included (Canada, England, Germany and Japan). The key differences are linked to the emergence of two new themes (Themes 6 and 7) that explain how structural issues across healthcare systems affect COPD care. This difference in the theme structure emphasises the importance of including countries with less resilient healthcare systems, such as Argentina, Mexico, and Russia (27). Through this inclusive approach, this research provides a better understanding of both shared and unique challenges in COPD care in UMICs and HICs.

The themes identified in this study are consistent with another publication that suggests the quality standards in COPD care (28). Four out of five quality standards suggested by a panel of clinical experts and patient representatives (timely diagnosis, patient awareness, availability of treatment options and adequate management of exacerbations) overlap with the themes reported here.

Kayyali et al. mapped and compared the COPD care delivery pathways in five European countries and, similarly to our findings, reported that most challenges were shared in all countries despite pathway variation (10). These included different manifestations of healthcare system fragmentation, which we also identified as a theme (Theme 3). Kayyali et al. emphasised the lack of a specialised role for pharmacists and informal carers (10), which the clinicians interviewed in our study did not report. However, this does not necessarily mean that these are not relevant issues in the countries we studied, but might rather reflect differences in the discussion guides.

### Interpretation of the findings

Clinicians in both HICs and UMICs face similar barriers, but the scale of the problem described in each theme varies (as outlined in Tables 4-10). Our results suggest UMICs experience a higher burden of COPD and associated challenges (29).

Clinicians’ observations revealed a notable difference in the role of exacerbations as drivers of COPD diagnosis between HICs and UMICs. In UMICs, clinicians reported that over 50% of COPD diagnoses are made following an exacerbation. In contrast, this was substantially lower in HICs (10-20%), which is consistent with other literature on exacerbations (30,31). These perspectives suggest that in UMICs, there might be an even greater delay in recognising and diagnosing COPD, which could impact timely intervention and disease management.

There were disparities in the perceived availability of services and treatment options for non-pharmacological therapy between HICs and UMICs. Clinicians in Spain, Australia, and Taiwan report better availability of non-pharmacological therapies than that reported by clinicians in the UMICs. However, even in these HICs, clinicians acknowledge that existing services are still inadequate to meet the needs of COPD patients. This emphasises the importance of improving non-pharmacological therapy options across all countries and is consistent with other publications (32,33).

Pulmonary rehabilitation was reportedly more accessible in Taiwan and Australia compared to Argentina, Mexico, and Russia, likely reflecting better infrastructure in HICs. Patient management programmes, inhaler technique education, and smoking cessation services were also reportedly more available in HICs. However, HICs and UMICs may face different challenges in these services (34).

Treatment availability and affordability reportedly pose greater challenges in UMICs (Argentina, Mexico, Taiwan) than in HICs (Australia, Spain). This indicates the importance of addressing affordability and expanding access to essential treatments and preventive measures in UMICs (35). Given that clinicians in Russia are deliberately misdiagnosing COPD patients with asthma so patients can get reimbursed, indicates that despite COPD’s high burden, it may not receive the same prioritisation and financial support as other non-communicable diseases (NCDs) (36). This also an issue in HICs.

There were notable differences between public and private healthcare provision in Mexico and Argentina, manifested as stark disparities in COPD care depending on the health insurance individuals can afford. The prominent role of private healthcare was not reported in other countries.

### Strengths and limitations

Interviews with clinicians from six countries facilitated a comprehensive analysis that provides a broader perspective on challenges in diverse healthcare settings. It also enabled us to compare and contrast barriers to the guideline-directed management of COPD in HICs and UMICs. To the best of our knowledge, this comparison has not been previously reported.

Including the graphic presentations of care pathways in the interviews facilitated meaningful discussions with clinicians, as they could see a comprehensive overview of COPD care in their respective countries when considering barriers and deviations from guidelines.

One limitation is that interviews occurred in different periods and by multiple researchers, which may have introduced inconsistencies in how questions were asked. Nevertheless, all researchers utilised the same discussion guides, minimising potential inconsistencies. Additionally, some interviews required interpretation and translation of the transcripts, which may have resulted in the loss of certain subtleties. However, since the analysis did not focus on nuances behind clinicians’ statements, this limitation is of lesser concern.

Lastly, when comparing the barriers between the countries, the differences may not be attributable to real variations in care provision but rather to the information the interviewee gave. It is possible some of their views were not expressed due to a lack of probing by the interviewer.

### Policy implications

The implications of our findings extend beyond this study. Policymakers can utilise the insights from the themes to identify areas where healthcare systems needs strengthening for COPD patients. Furthermore, given that most barriers are similar across healthcare settings, there are opportunities for concerted efforts in COPD care through international policy initiatives. While there are many overarching strategies for NCDs generally (37), there is no coordinated strategy to encourage countries to prioritise the prevention and management of COPD (e.g,, improved diagnosis through better use of spirometry and guideline-adherent therapy), which is one of the main NCDs driving up global mortality and morbidity rates (38). Similar to other NCDs, the findings of this research can inform efforts to develop funded national respiratory strategies (39,40).

Policymakers can leverage the visual presentation of the Evidenced Care Pathways to facilitate discussions, align goals, and encourage interdisciplinary cooperation, leading to better coordination of care, reduced fragmentation, and improved patient outcomes (41).

For future research, exploring approaches that further unify and systematise care pathway mapping would be valuable, ensuring academic rigour while informing policy action. The replication of the Evidenced Care Pathways in other countries could be a starting point.

## Conclusion

HICs and UMICs differ regarding awareness levels among patients and HCPs, primary care involvement, spirometry access, and specialised care availability. While both face challenges related to fragmented healthcare systems, guideline adherence, and COPD exacerbation management, UMICs struggle with additional issues stemming from limited resources and healthcare infrastructure.

Most barriers to COPD care are similar across HICs and UMICs, indicating that COPD remains a persistent challenge. While unique issues need to be addressed through tailored approaches, there are missed opportunities to implement international respiratory strategies that encourage countries to take measures to manage COPD adequately. Alongside efforts at the healthcare system level, there is a need for political prioritisation of COPD to ensure it is allocated the necessary resources, even in HICs.

## Data Availability

All data produced in the present study are confidential and cannot be shared; a selection on anonymised data are available in the manuscript.

## Acknowledgements

We want to thank all participants who contributed valuable insight to depict current care for COPD. Many thanks go to Alice Sinclair for coding the transcripts and Mathilde Vankelegom, Nuri Ahmed, Paddy Goddard-Watts, and Aleix Rowlandson for leading some interviews. Lastly, we would like to express our gratitude to Marta Wanat for providing her qualitative expertise to improve the design and reporting of this research.

## Author contributions

All authors made a significant contribution to the work reported. RGA, KT, and EA were involved in the conception, study design, and execution. KPS was involved with the acquisition of data. OS and RGA performed the analysis and interpretation. All coauthors took part in drafting, revising or critically reviewing the article; gave final approval of the version to be published; agreed on the journal to which the article has been submitted; and agree to be accountable for all aspects of this work.

