## Supplemental file for "Mapping the common barriers to optimal COPD care in high and middle-income countries: qualitative perspectives from clinicians"

### Supplement 1 – Search strategy

PubMed and grey literature (conference abstracts, HTA reports, publicly available national databases reporting on COPD outcomes) were searched, and reference lists of relevant papers were reviewed.

#### Argentina

Search terms:

- COPD OR chronic obstructive pulmonary disease AND [Argentina]
- “COPD patient pathway” AND [Argentina]
- COPD OR chronic obstructive pulmonary disease AND “clinical care” AND [Argentina]
- COPD OR chronic obstructive pulmonary disease AND management AND [Argentina]
- COPD OR chronic obstructive pulmonary disease AND economic AND [Argentina]

#### Australia

- COPD OR chronic obstructive pulmonary disease AND [Australia]
- “COPD patient pathway” AND [Australia]
- COPD OR chronic obstructive pulmonary disease AND “clinical care” AND [Australia]
- COPD OR chronic obstructive pulmonary disease AND management AND [Australia]
- COPD OR chronic obstructive pulmonary disease AND economic AND [Australia]

#### Mexico

- COPD OR chronic obstructive pulmonary disease AND [Mexico]
- “COPD patient pathway” AND [Mexico]
- COPD OR chronic obstructive pulmonary disease AND “clinical care” AND [Mexico]
- COPD OR chronic obstructive pulmonary disease AND management AND [Mexico]
- COPD OR chronic obstructive pulmonary disease AND economic AND [Mexico]

#### Spain

- COPD OR chronic obstructive pulmonary disease AND [Spain]
- “COPD patient pathway” AND [Spain]
- COPD OR chronic obstructive pulmonary disease AND “clinical care” AND [Spain]
- COPD OR chronic obstructive pulmonary disease AND management AND [Spain]
- COPD OR chronic obstructive pulmonary disease AND economic AND [Spain]

#### Russia

- COPD OR chronic obstructive pulmonary disease AND [Russia]
- “COPD patient pathway” AND [Russia]
- COPD OR chronic obstructive pulmonary disease AND “clinical care” AND [Russia]
- COPD OR chronic obstructive pulmonary disease AND management AND [Russia]
- COPD OR chronic obstructive pulmonary disease AND economic AND [Russia]

#### Taiwan

- COPD OR chronic obstructive pulmonary disease AND [Taiwan]
- “COPD patient pathway” AND [Taiwan]
- COPD OR chronic obstructive pulmonary disease AND “clinical care” AND [Taiwan]
- COPD OR chronic obstructive pulmonary disease AND management AND [Taiwan]
- COPD OR chronic obstructive pulmonary disease AND economic AND [Taiwan]

### Supplement 2 – Structure of a discussion guide used during the clinician interviews

Semi-structured interviews were carried out using the interview topic guide presented below. The interviews were conducted in an iterative manner, with the questions in the guide being modified to encompass emerging themes and enhance the overall direction. It should be noted that the questions provided in the topic guide are not exhaustive, meaning that additional follow-up questions were posed based on the participants' responses. Furthermore, it is important to acknowledge that not all questions were asked in every country.

**Introduction to the project - 10 mins**

Background on the project:

*The aim is to develop a representative version of the overall care structure and more detailed or micro-level pathways for a given disease area, in this case, COPD.*

*This is then populated with existing data, such as the number of patients and time taken between steps, rather than data generated through modelling or by making assumptions. This structure and data will be used to help identify potential blocks or barriers in the pathway – which can then be raised and discussed with, in this case, policymakers, as well as for internal strategy purposes.*

*Our role is to develop representative pathways for COPD across six countries, [country] being one, and then populate it with published data.*

Explain the expert role:

*“To take a critical view of the pathways so far developed for [country], helping us shape, correct and fill any gaps – based on their experience (even if that is a partial view – we will be talking to other experts to achieve balance)*

*To answer any questions derived from our research (to the best of their knowledge).*

*If possible/where relevant, comment on the (essential elements of) data we have sourced, or perhaps provide direction for our further research.”*

- Field any questions from them, as appropriate

**Context setting: expert’s role and background/experience in COPD – 10 mins**

- Please tell us about a relevant background in respiratory medicine (what roles, who is treating/what researching (as appropriate), how long etc.)
- And in COPD particularly?
- Reflect on the main issues, from their point of view, in the status or provision of treatment of COPD in [country]
  - *(this is not intended to replace or duplicate the discussion below but more to cue up their perspective and capture the most significant issues so that we can gear the questioning accordingly*)
  - What impact do you think COVID-19 will have on the future pathway and management of COPD in [country]?

**The main phase of discussion: running through the pathway(s) as it has been developed so far – 10 mins**

- Explain how we intend to present the pathways and roughly how we expect the discussion to work:

*“We’ll be using PowerPoint to show the pathways. Hopefully, the structure/position of the pathways will be clear as we move through the PowerPoint. I’ll be taking us through them, hopefully presenting you with a sufficient overview of each, then going through (the areas of most interest) step by step.*

*Please tell me to move around and zoom in and out as and when you want to.*

*As the project has progressed, especially now that the [country] pathway is formatted nicely and looks ‘designed’, please don’t assume this means it is complete or correct. We have found that sometimes people think the pathway tends to feel more concrete, which can lead to more confirmatory findings. They still very well could be wrong, so please view them critically!”*

- We are likely to present the pathways in this order/with this approach:
  - An overview of the macro level structure of care (Table of Contents) for [country]
    - Give an overview of how the pathway is established and how it’s been broken down
    - (*We need to ensure we mention that a patient sees the specific details such as health services used, or HCP will be discussed in the micro pathways rather than the macro)*
    - What do they see as the main issues or blocks in terms of diagnosing/treating COPD?
      - Discuss some of the underlying causes of this
      - What data do we have to support this?

**More detailed run-through – moving to the micro-level pathways – 55 mins**

*[Note: questions will also be targeted according to what the countries are interested in]*

- Explain to them the order we are considering exploring at the micro-level in [as appropriate to the country] order.
- Next, discuss the approach to the micro-level pathways session. We will have control of the screen and can move through the pathways at our own pace. As we move through the pathway, we will ask any particular questions at each step. Let them know that we can at any time come out of the micro-level pathway and reflect on any macro-structural implications, should this be needed (e.g. an extra step, renaming a step)
- Check with them whether they are comfortable with this approach
- Then check if there is anything they would like to discuss first/prioritise (e.g. because it would have implications on other aspects of the pathway).

*Questions to ask*

- Specifics we may want to ask at this point:
  - Who is giving care?
  - Patient flows – do these look about right
    - Directionally
    - Proportionately
  - Where do you think COVID will have the most impact?
  - What are the timelines between each step?
  - Are there any points in the pathway where care delivery varies hugely from the recommendations?
  - Refer to [country-specific Excel doc or comments in the pathway] for specific questions on the micro-level pathway.

**Round up: summary of main considerations to take forward** **– 5 mins**

- Summarising the main issues/barriers to better COPD treatment in [country] and the main difference/details they have identified
- Unanswered questions for Aquarius to go and research), any other thoughts.
